# Absence of *Glutathione S-Transferase Theta 1* Gene is Significantly Associated with Breast Cancer Susceptibility in Pakistani Population and Poor Overall Survival in Breast Cancer Patients: A Case-Control and Case Series Analysis

**DOI:** 10.1101/2021.02.18.21252044

**Authors:** Sadia Ajaz, Sani-e-Zehra Zaidi, Saleema Mehboob Ali, Aisha Siddiqa, Muhammad Ali Memon, Sadaf Firasat, Aiysha Abid, Shagufta Khaliq

## Abstract

**Purpose:** Deletion of <u>G</u>lutathione <u>S</u>-<u>T</u>ransferase <u>T</u>heta <u>1</u> (GSTT1) encoding gene is implicated in breast cancer susceptibility, clinical outcomes, and survival. Contradictory results have been reported in different studies. The present investigation evaluated *GSTT1*-absent genotype for its’ contribution to breast cancer risk in Pakistani population and specific clinical outcomes in breast tumours.

**Methods:** A prospective study comprising case-control analysis and case series analysis components was designed. Peripheral blood samples were collected from enrolled participants. After DNA extraction, *GSTT1 g*enotyping was carried out by a multiplex PCR with *β-globin* as an amplification control. Association evaluation of *GSTT1* genotypes with breast cancer risk, specific tumour characteristics, and survival was the primary endpoint.

**Results:** A total of 264 participants were enrolled in the molecular investigation (3 institutions). The study included 121 primary breast cancer patients as cases and 143 age-matched female subject, with no history of any cancer, as controls. A significant genetic association between *GSTT1*-absent genotype and breast cancer susceptibility (*p*-value: 0.003; OR: 2.13; 95% CI: 1.08-4.29) is reported. The case-series analysis showed lack of association of *GSTT1* genotypes with tumour stage (*p*-value: 0.12), grade (*p*-value: 0.32), and size (*p*-value: 0.07). The survival analysis revealed that *GSTT1*-absent genotype cases had a statistically significant shorter overall survival (OS) than those with *GSTT1*-present genotype cases (mean OS: 23 months vs 33 months). The HR (95% CI) for OS in patients carrying *GSTT1*-absent genotype was 8.13 (2.91-22.96) when compared with *GSTT1*-present genotype.

**Conclusions:** The present study is the first report of an independent, population-oriented significant genetic association between *GSTT1*-absent genotype and breast cancer susceptibility as well as OS in breast cancer cases. Upon further validation, *GSTT1* variation may serve as a marker for devising better and population-specific strategies for screening and treatment in breast cancer management.

## Introduction

Glutathione is present in all living cells. Physiologically, it performs three important functions: protection of thiol groups in proteins from oxidation, intracellular redox buffering, storage for sulphur-containing cysteine. These functions are dependent upon the catalysis by <u>G</u>lutathione <u>S</u>-<u>T</u>ransferases, E.C. 2.5.1.18. Consequently, GSTs play a major role in detoxification of potent endogenous and exogenous carcinogens [1]. These enzymes constitute a superfamily of isoenzymes including <u>GST</u>-theta <u>1</u>. *GSTT1* gene is located on chromosome 22q11.2. It encodes the enzyme, which is involved in conjugation of reduced glutathione to various electrophiles and hydrophobic compounds [2]. Ultimately, toxic substrates may be removed from the body.

The absence of *GSTT1* gene, also known as homozygous deletion or null genotype and herein referred to as *GSTT1*-absent, has been reported with varying frequencies in different populations [3]. The carriers of *GSTT1*-absent genotype are unable to metabolize specific mutagenic carcinogens [4]. The deletion has been correlated with ovarian, bladder, colon, oral, lung and pediatric cancers among different populations [5-10]. It is a candidate genetic markers for cancer risk, prognosis, and treatment response. The independent contribution of *GSTT1* null genotype to breast cancer susceptibility, tumour characteristics, and response to prescribed regimens remains inconclusive in different populations across the world [11-13].

In Pakistan, the age-standardized rate (ASR) of the female breast cancer incidence is among the highest in Asia (43.9 per 100,000), whereas the mortality rate is one of the highest in the world (23.2 per 100,000) [14, 15].

Two previous studies from Pakistan [16, 17] report an independent lack of association between absence of *GSTT1* gene and breast cancer susceptibility. Both the studies were published from the Punjab area. Furthermore, the small sample size, and conflicting frequencies in controls: 18.7% [16] vs 31.4% (erroneously reported as 16% in the study) [17] limit the applicability of drawn conclusions.

The prospective observational molecular study was designed based on the biological plausibility of *GSTT1* deletion in carcinogenesis. It addresses the paucity and contradiction in the available data from a region that has frequent and aggressive breast tumors. The first component of the study, the case-control analysis, evaluated the contribution of *GSTT1* gene in breast cancer risk. Simultaneously, the second part comprising case series analysis investigated the contribution of *GSTT1* genotypes in selected tumour characteristics and breast cancer survival after standard treatment.

## Materials and methods

### Study design and participant enrollment

The overall study schema is shown in Fig 1.

**Fig. 1.**
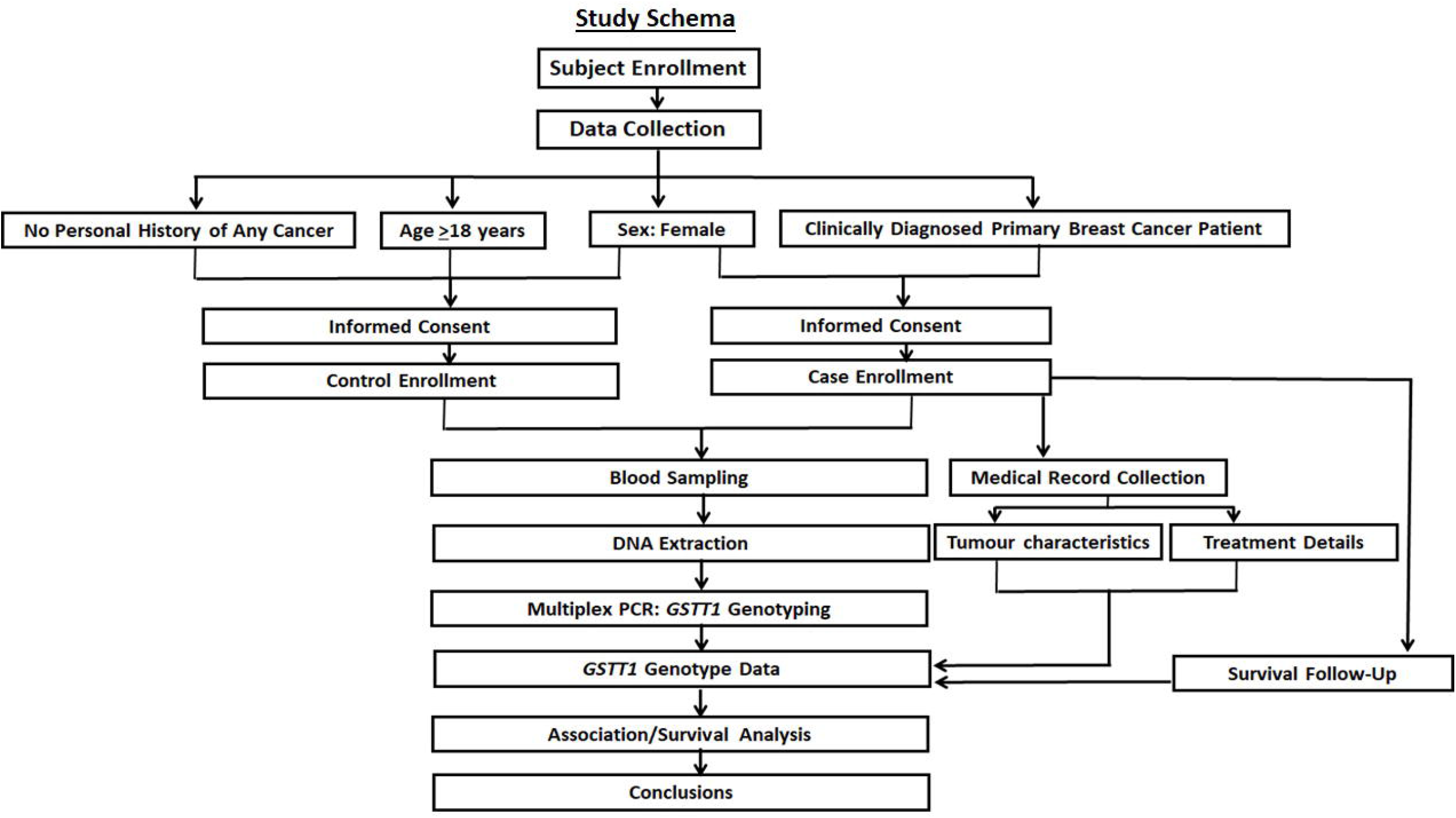
**The study schema. The prospective recruitment of cases with age-matched female controls is shown along with the collection of specified molecular, clinical, and survival data. The objective was to investigate contribution of *GSTT1* variation in breast cancer risk, tumour characteristics and survival after standard treatment.**

The patients were recruited from <u>A</u>tomic <u>E</u>nergy <u>M</u>edical <u>C</u>entre (AEMC), <u>J</u>innah <u>P</u>ostgraduate <u>M</u>edical <u>C</u>entre (JPMC), Karachi, Pakistan. The cases were clinically diagnosed primary breast cancer patients. The cases underwent radiotherapy at the aforementioned participating institution following chemotherapy and surgery, which were carried out at hospitals other than AEMC. The details of control enrolment have been published elsewhere [18], with the modification of inclusion of only samples from age-matched (≥ 18 years), female participants in the present study. All the subjects were recruited at Karachi, Pakistan and therefore, the distribution of ethnicities was the same in cases and controls (Sindhi, a self-defined Urdu-speaking ethnicity, Pathan, and Punjabi were the main ethnic groups). The research involved human participants and followed the provisions of Declaration of Helsinki and its amendments. Research protocols were approved by the independent Ethics Review Committees of all the relevant institutions. The study follows the <u>re</u>porting recommendations for tumor <u>mark</u>er prognostic studies (REMARK) [19, 20] (Supplementary Table 1).

### Data collection

Information regarding age was recorded from all the participants. In patients, tumor node metastasis (TNM) staging and histological grading were carried out according to the Union Internationale Contre le Cancer (UICC) recommendations [21]. Data pertaining to tumour characteristics (tumour stage, grade, and size) and histology were obtained from the patients’ hospital medical files. The information for the included parameters was documented in majority of the hospital records, unlike other parameters associated with breast cancers, for e.g. hormonal status, and Bloom Richardson grading system etc. Three-year survival data were collected through telephonic follow-up. The missing information is due to: (i) return of patients to their own towns/villages after treatment at Karachi; (ii) erroneous contact information; (iii) and no response.

### Sample collection and DNA extraction

All the participants volunteered 8-10ml of venous blood sample, which was collected in ACD-coated vacutainers (BD Vacutainer^®^ BD Franklin Lakes NJ USA). Samples from the cases were collected at the time of radiotherapy, post-mastectomy and chemotherapy treatment. The blood samples were either processed immediately or stored at 4^°^C until DNA extraction.

DNA was extracted from the white blood cells according to the standard phenol-chloroform method [22]. It was quantified spectrophotometrically (Beckman Coulter(tm) DU^®^ 530). The quality control cut-off for 260/280 ratio was between 1.7-1.99. DNA quality was also analyzed by 0.7% agarose gel electrophoresis followed by UV visualization using a gel imaging system (Azure c300^®^ biosystems). No fragmentation or smearing was observed in any of the samples (Fig 2).

**Fig 2.**
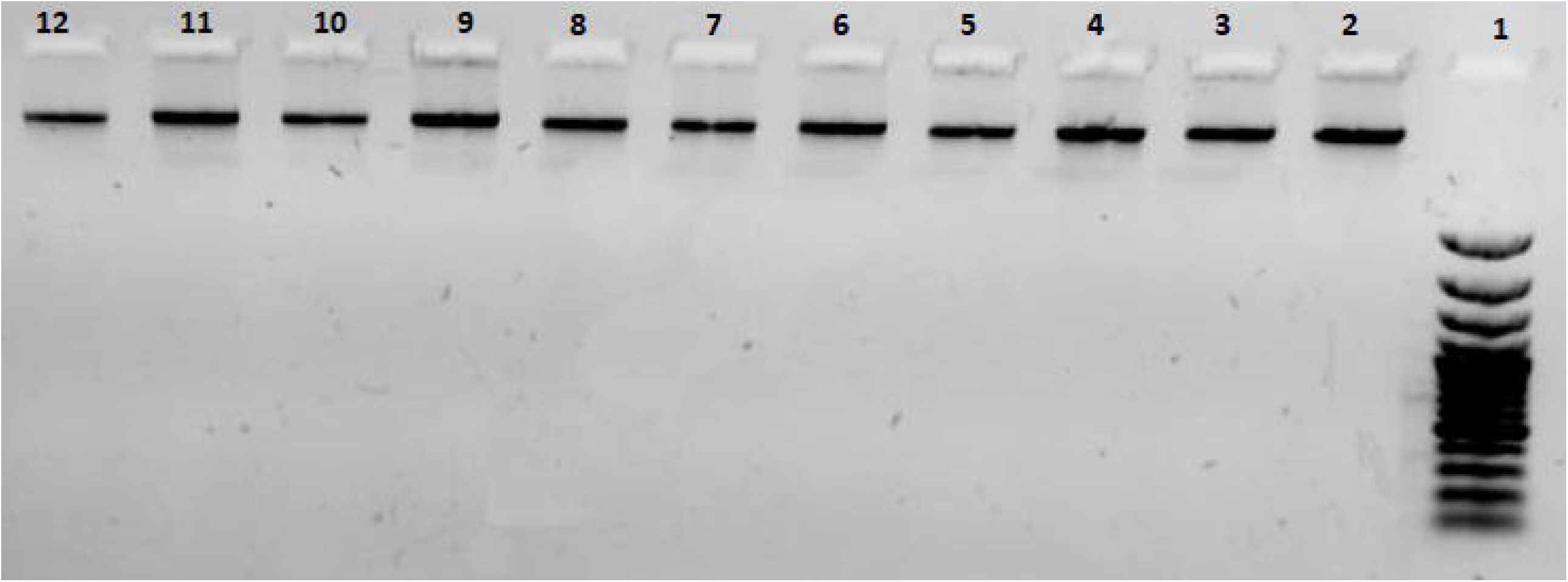
**A representative 0.7% agarose gel used for quality control of extracted DNA samples. Lane 1: 100bp DNA ladder; lanes 2-12: DNA samples**

The working dilutions for experiments were prepared at room temperature and stored at 4^°^C. The stock DNA samples were stored at −20^°^C.

### Genotyping

#### Cases

*GSTT1* genotyping was carried out by a multiplex polymerase chain reaction (PCR) with *β-globin* as an amplification control. The primer sequences have been published earlier [17]. PCR was carried out with *Taq* DNA polymerase kit (Thermofisher Scientific Inc.). Total PCR reaction mix (10µl) consisted of 1X PCR buffer, 0.9mM MgCl_2_, 0.5mM dNTPs, 1.5U/µl *Taq* polymerase, 1.8µM primers each for *GSTT1* and *β-globin* genes, and 70ng DNA. The PCR conditions were: initial denaturation at 94°C for 5 minutes, followed by 40 cycles of: 94°C for 45 seconds, annealing at 60°C for 45 seconds, and extension at 72°C for 45 seconds. Final extension was carried at 72°C for 5 minutes. Amplicons were analyzed under UV on 2% agarose gel, which was stained with ethidium bromide. A fragment of 473bp indicated *GSTT1*-present genotype. *GSTT1*-absent genotype did not show this amplification. The amplification of β-globin gene served as the control for successful PCR. A negative control was included in all the genotyping experiments (Fig. 3).

**Fig. 3.**
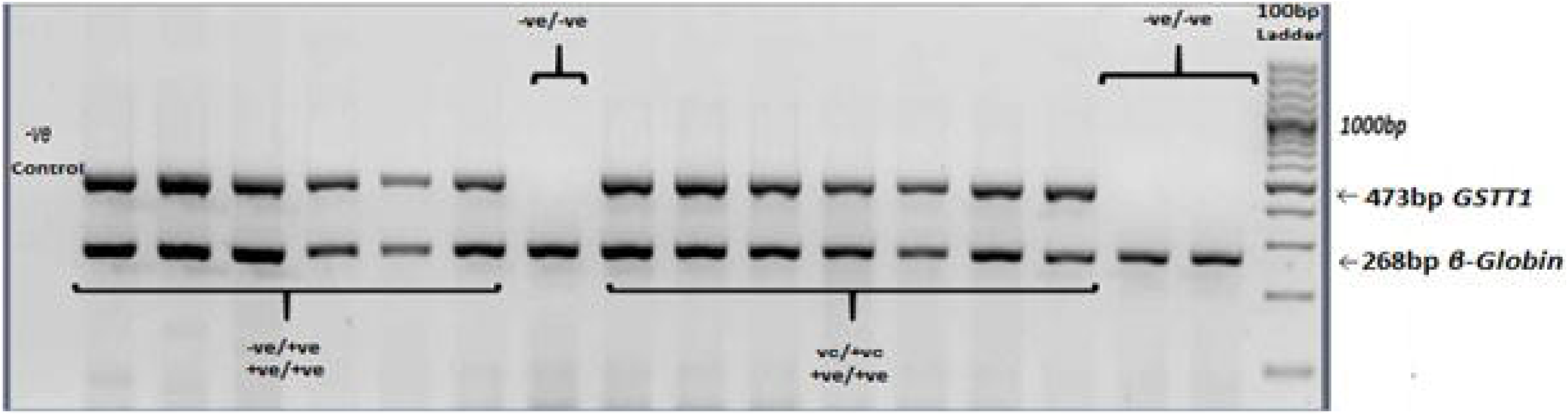
***GSTT1* genotyping. Agarose gel electrophoresis (2%) of multiplex PCR amplified products: *GSTT1*-absent (–ve/-ve) genotype did not show amplification of a 473bp fragment. *β-globin* gene was included as an amplification control.**

The results of genotyping were confirmed by double blind evaluation, inclusion of replicates, and negative controls.

#### Controls

The genotyping in controls has been described earlier [18]. It differentiates between homozygous *GSTT1-*present, heterozygous *GSTT1-*present/absent, and homozygous *GSTT1-* absent. In the final observation, *GSTT1*-present allele was determined by an amplicon of 466bp, while *GSTT*-absent allele was identified by an amplicon of 1,460bp.

### Treatment of breast cancer patients

All the participating cases underwent mastectomy, adjuvant chemotherapy, and/or radiotherapy. Before the start of chemotherapy, echocardiography was done to assess cardiac function (ejection fraction cut-off for start of doxorubicin based chemo was >55%). A test dose of docitaxel was given to rule out hypersensitivity before 1^st^ cycle. Neurological assessment was done during taxane (paclitaxel) cycles. Chemotherapy related toxicities were assessed after every cycle according to <u>C</u>ommon <u>T</u>oxicity <u>C</u>riteria of the <u>N</u>ational <u>C</u>ancer Institute (NCI-CTC, version 2.0) [23]. The “severe toxicity” was defined as hematological or gastrointestinal toxicity of grade 3–4.

The followed chemotherapy regimen was: Adriamycin-Cyclophosphamide x4 followed by taxane x4 (docitaxel or paclitaxel): Doxorubicin 60mg/m^2^ on day 1, cyclophosphamide 600mg on day 1, paclitaxel 175mg/m^2^ on day 1 OR docitaxil 100mg/m^2^ on day 1. Repeated every 3 weeks.

Complete blood count, liver function test, and renal function test were carried out to assess the treatment response.

### Statistical analysis

The allele distribution for *GSTT1* polymorphism in controls was assessed for Hardy-Weinberg equilibrium [24]. The statistical tests for association analysis were carried out by using Statistical Package for Social Science (SPSS) for Windows v.19.0 (SPSS, Inc., Chicago, Illinois, USA) and online OpenEpi software [25]. In case-control investigations, data for *GSTT1* genotype was obtained for all the participants, except two cases where no amplification was recorded. The age-matching between cases and controls was analyzed by Student’s t-test for independent samples with the assumption of unequal variances. To achieve 80% power at a two-sided level of significance, various odds ratios (OR) of genetic risk due to *GSTT1* polymorphism for breast cancers were calculated. Accrual of 260 participants (matched cases and controls) allows the identification of OR≥2 for *GSTT1* variation with the *GSTT1*-absent frequency of 0.24 (the median value of the reported prevalence in controls from Pakistan [16-18, 26-37] was used for the calculations [38]).

The missing information for case series analysis is itemized in the relevant results section. The primary objective was the investigation of *GSTT1* polymorphism association/s with breast cancer susceptibility, the selected clinical parameters, and survival. The data were assessed by Pearson χ^2^ test. The OR were tabulated with 95% confidence interval (95% CI) to evaluate the strength of the associations. The overall survival (OS) and hazard ratios (HR) with 95% CI were assessed by Kaplan-Meier method using MedCalc software v.19.2.6 [39, 40]. In all the statistical tests, p-values <0.05 were considered to be significant.

## Results

### Participants’ information and clinical data of patients

Total number of patients diagnosed with primary breast cancer disease was 121, whereas the total number of age- and gender-matched controls was 143. Characteristics of the 264 participants included in the study are presented in Table 1.

**Table 1.** Participant information and clinico-pathological data of breast cancer patients.

| Sr. No. | Characteristic | Value | *p-value* |
| --- | --- | --- | --- |
| 1. | No. of participants (cases/controls) | 264 (121/143) | N/A |
| 2. | Mean Age [cases/controls: years±standard error of mean | 44.48±0.95/45.62±0.58 | 0.306 |
| 3. | Tumour Stage (*n* *= 84) |  |  |
|  | I and II | I: 3 (4%) and II: 30 (36%) | 0.024** |
|  | III and IV | III: 46 (55%) and IV: 5 (5%) |  |
| 4. | Tumour Size (*n* *=102) |  |  |
|  | < 2cm | 14 (14) |  |
|  | 2-5cm | 58 (57) | <0.01** |
|  | >5cm | 30(29) | >0.05 |
| 5. | Tumour grade (*n* *=107) |  |  |
|  | G1 and G2 | G1: 1 (1%) and G2: 46 (43) | >0.05 |
|  | G3 and G4 | G3: 58 (54%) and G4: 2 (2%) |  |
| 6. | Treatment Response (*n* *=97) |  |  |
|  | Positive | 64 (66%) | <0.01** |
|  | Negative (relapse and/or death) | 33 (34%) |  |
| 7. | 3-Year Survival (*n* *=97) |  |  |
|  | Alive | 71 (73%) | <0.01** |
|  | Expired | 26 (27%) |  |

\*available data from 121 patients (missing data has been explained in the methodology section);
\*\*statistically significant

Mean age of the patients was 44.48±0.95 years, whereas for the controls, the mean age was 45.62±0.58 years. All patients presented with invasive ductal carcinoma (IDC) of the breast. Majority of the patients had advanced tumor stage (stages III and IV; *p-*value: 0.024**), tumour size of >2cm (*p*-value: <0.01**), and high tumour grade (grades 3 and 4; *p*-value: >0.05).

### Association between *GSTT1* polymorphism and breast cancer risk

Allelic and genotypic frequencies of *GSTT1* polymorphism in controls are shown in Table 2. The proportions were in Hardy-Weinberg equilibrium.

**Table 2.** **Distribution of *GSTT1* genotypes and allele frequencies (with standard errors) in age- and gender-matched controls. Assessment of HWE test in controls**.

| ***GSTT1* Polymorphism** | **Controls (n = 143)** |
| --- | --- |
| *Genotypes* |  |
| *GSTT1*-present/ *GSTT1*-present | 66 (46%) |
| *GSTT1*-present/ *GSTT1*-absent | 61 (43%) |
| *GSTT1*-absent/ *GSTT1*-absent | 16 (11%) |
| *Allele Frequencies* |  |
| p[*GSTT1*-present] | 0.67±0.042 |
| q[*GSTT1*-absent] | 0.33±0.042 |
| *Hardy-Weinberg Equilibrium (HWE) Test* |  |
| $\chi^2$ | 0.11 |
| *P*-value | NS (0.74) |

Associations between the *GSTT1* genotypes and breast cancer susceptibility are presented in Table 3.

**Table 3.** **Distribution of *GSTT1* genotypes in controls, breast cancer patients, and the association analysis with breast cancer risk**.

| *GSTT1* Polymorphism | Controls<br>(n = 143) | Breast Cancer Patients<br>(n = 118<sup>¶</sup>) | $\chi^2$ Test (*p*-value) | OR (95% CI) |
| --- | --- | --- | --- | --- |
| *GSTT1*-absent/ *GSTT1*-absent | 16 (11%) | 25 (21%) | $\chi^2= 4.88$;<br>*p*=0.03** | 2.13 (1.08-4.29)** |
| *GSTT1*-present/ *GSTT1*-present &<br>*GSTT1*-present/ *GSTT1*-absent | 127 (89%) | 93 (79%) |  |  |

<sup>¶</sup>Genotype could not be determined for three samples; \*\*statistically significant

The comparison of *GSTT1*-present genotype with *GSTT1*-absent genotype in cases and controls revealed that *GSTT1*-absent genotype was significantly associated with risk for breast cancers (*p*-value: 0.003). The OR were 2.13 (95% CI: 1.08 - 4.29).

### Association between *GSTT1* polymorphism and specific tumour characteristics

In case series analysis, the present study does not report any statistically significant association between the absence of *GSTT1* gene and the studied tumour characteristics, i.e., stage, grade, and size. The *p-*values were 0.12, 0.32, and 0.07, respectively.

### Association of the *GSTT1* polymorphism and OS in breast cancer patients

As shown in Table 4 and Fig 4, *GSTT1*-present carriers had 10 months’ longer survival (mean OS: 33 months; 95% CI: 30.96-34.65) than those with *GSTT1*-absent genotype (mean OS: 23 months; 95% CI 17.90-28.59); *p*-value: 0.0001. The HR with 95% CI for OS in patients carrying *GSTT1*-absent genotype was 8.13 (2.91-22.96) with *GSTT1*-present genotype as the reference variable.

**Table 4.**
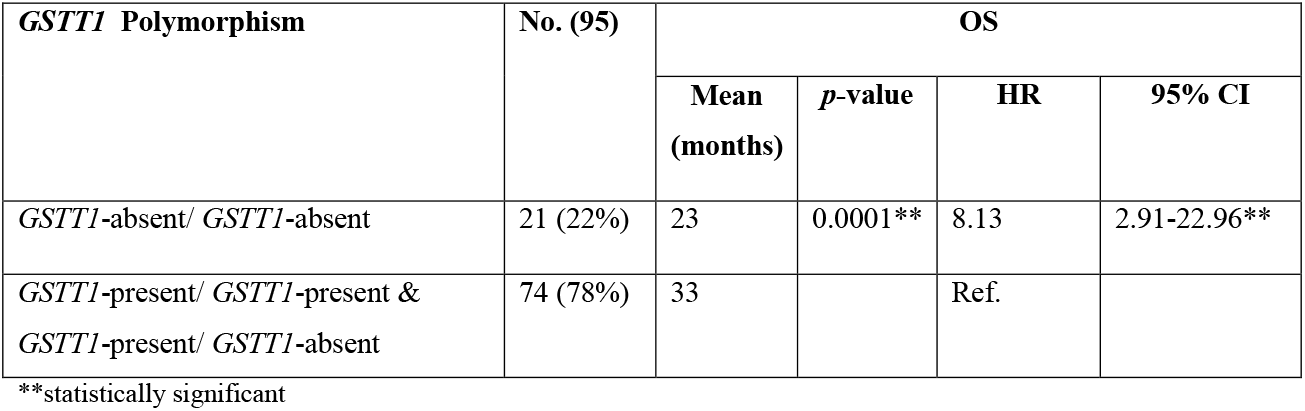
Associations between *GSTT1* genotypes and overall survival (OS)

**Fig 4.**
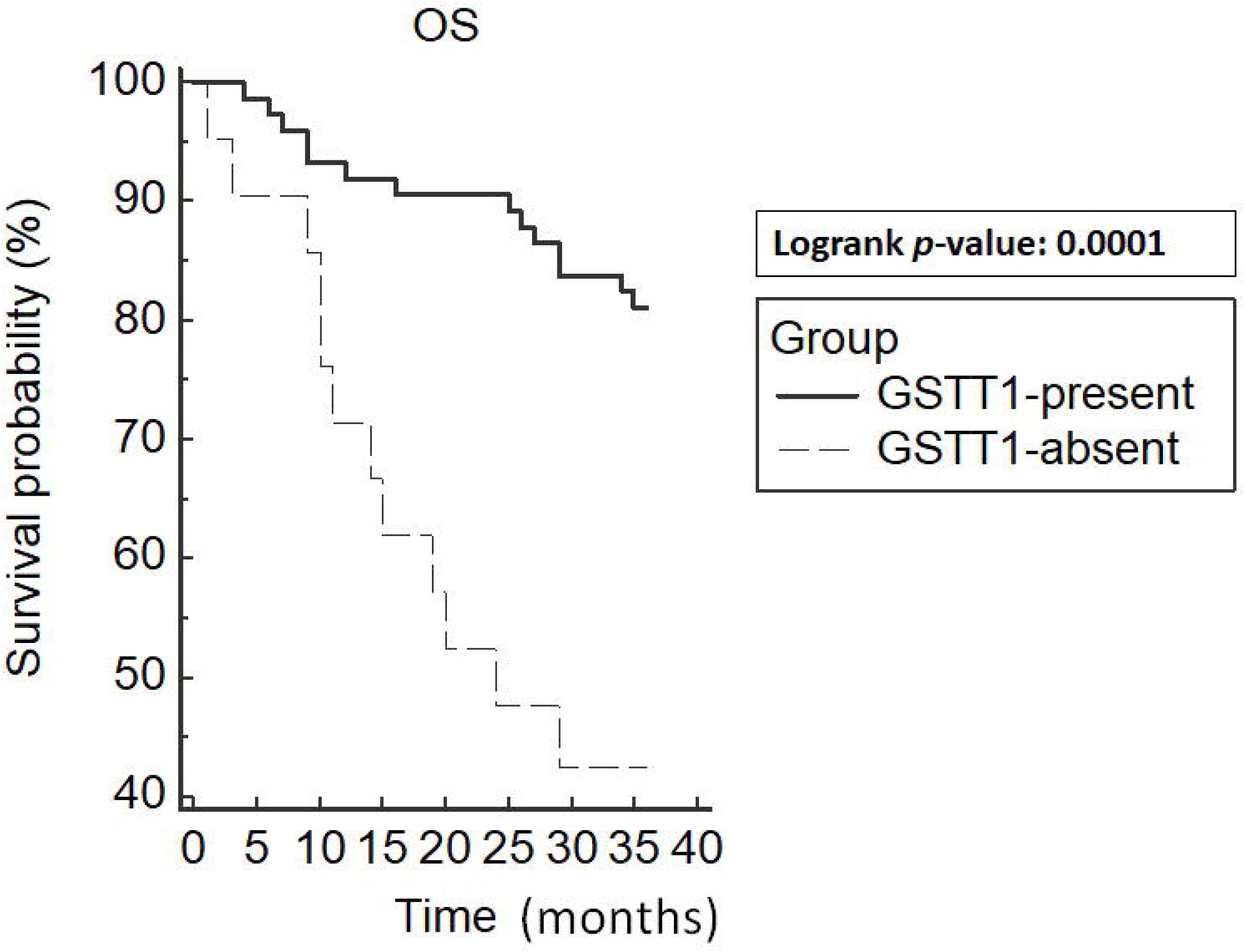
**Kaplan-Meier curve demonstrating the overall survival (OS) based on genotypes of *GSTT1*. The mean OS was 33 months (95% CI: 30.**96-34**.**65) in GSTT1-present genotype carriers and 23 months (95% CI: 17**.**90-28**.**59) in *GSTT1*-absent genotype carriers; *p*-value: 0**.**0001**.**

## Discussion

In the present study, we evaluated the association of *GSTT1* genotypes with breast cancer-related parameters. We report a *GSTT1*-absent frequency of 11% in the controls, which is in agreement with couple of earlier studies published from the region, one from Islamabad [33], and the other from Karachi [18]. In this study, we report a significant association of *GSTT1*-absent genotype with increased breast cancer risk in a representative sample from Pakistani population. The OR were 2.13 (95% CI: 1.08 – 4.29). We also report a significant difference in the survival duration between *GSTT1*-present and *GSTT1-*absent carriers: mean OS_*GSTT1*-present_: 33 months (95% CI: 30.96-34.65) vs mean OS_*GSTT1*-absent_: 23 months (95% CI: 17.90-28.59). The present analysis is the first report of such population-specific associations between *GSTT1* genotypes and specific factors associated with breast cancers.

The incidence of breast cancers varies across the globe. The highest estimated age-standardized incidence rates are reported from Belgium (113.2 per 100,000), while the lowest are reported from Bhutan (5.0 per 100.000). Furthermore, the highest estimated age-standardized mortality rates are reported from Fiji (36.9 per 100,000) while the lowest are reported from Bhutan (2.7 per 100,000) [15]. The known risk factors such as age, family history, different reproductive parameters, and obesity account for only one-third of the risk for breast cancers [11, 41]. In addition, the reason(s) for high mortality rates across certain populations need to be determined [42-44].

It is likely that a number of genes are involved, with the possibility of gene-environment interactions, in breast cancer etiology, progression, and response to treatment [45]. The quantitative contributions of such genes remain to be delineated across different populations and regions.

A proposed mechanism of carcinogenesis due to loss of function of GSTT1 isoenzyme is shown in Fig 5. The exogenous and endogenous carcinogens are not metabolized to non-toxic components. Consequently, tumourigenesis and/or tumour progression are likely to occur [4]. In addition, chemotherapeutic agents may also be metabolized by the pathways involving GSTT1, rendering the patients with *GSTT1*-present genotype irresponsive to either therapy or specific doses of therapy.

**Fig 5.**
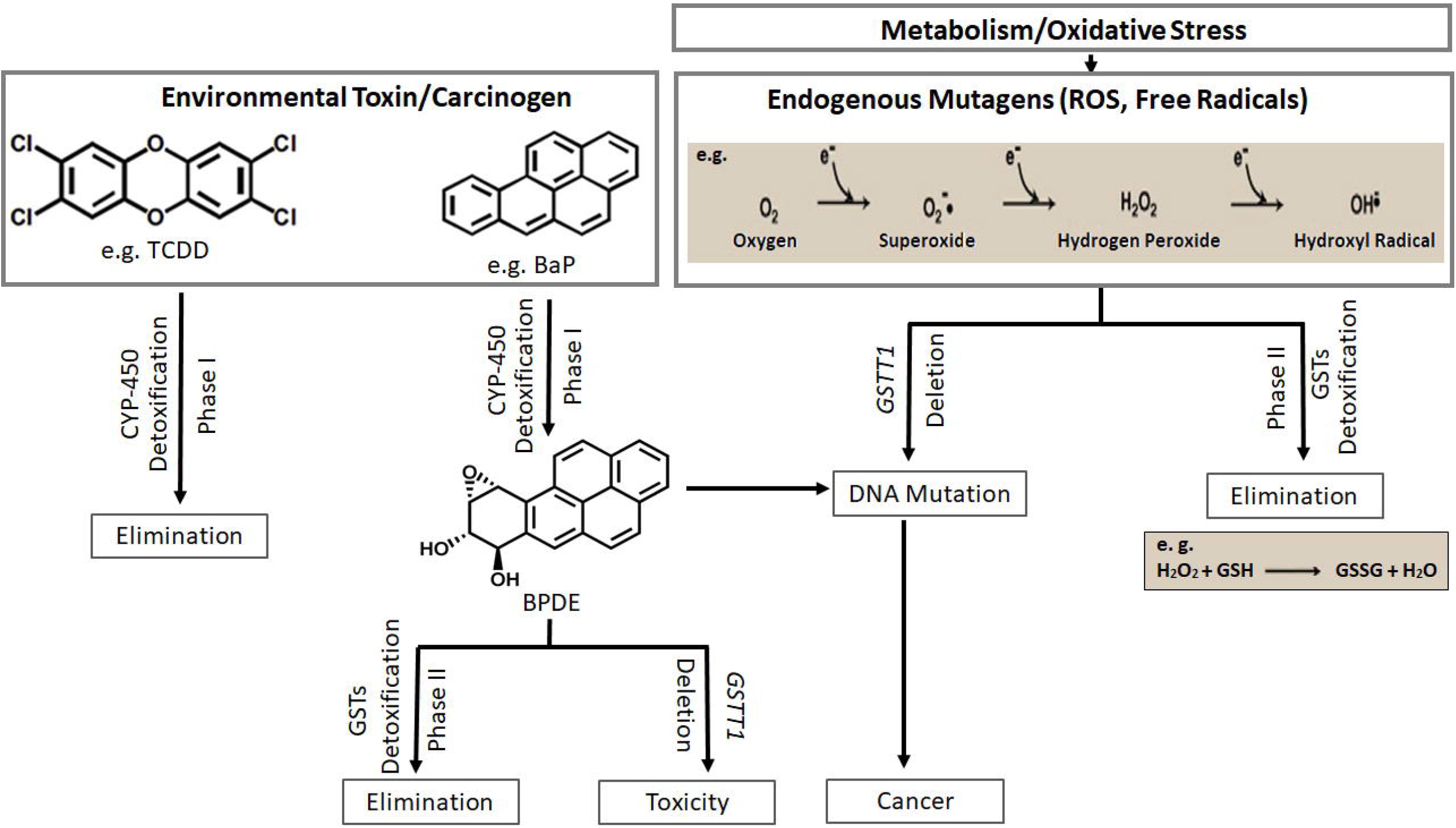
**Xeno- and endo-biotic carcinogen metabolism through two pathway system (phase I and phase II) is shown. Deletion of *GSTT1* leads to toxicity and carcinogenesis (TCDD: 2,**3**,**7**,**8-Tetrachlorodibenzodioxin; BaP: Benzo[a]pyrene; BPDE: Benzo(a)pyrene diolepoxide; ROS: Reactive Oxygen Species)****

Among different populations, the loss-of-function polymorphism in *GSTT1* encoding gene occurs with varying frequencies [46]. Null genotype is correlated with vulnerability to cancers, tumour characteristics, and differences in treatment response [1, 47].

The examples of low and high frequencies of *GSTT1*-absent genotype across different global regions are listed in Table 5.

**Table 5.**
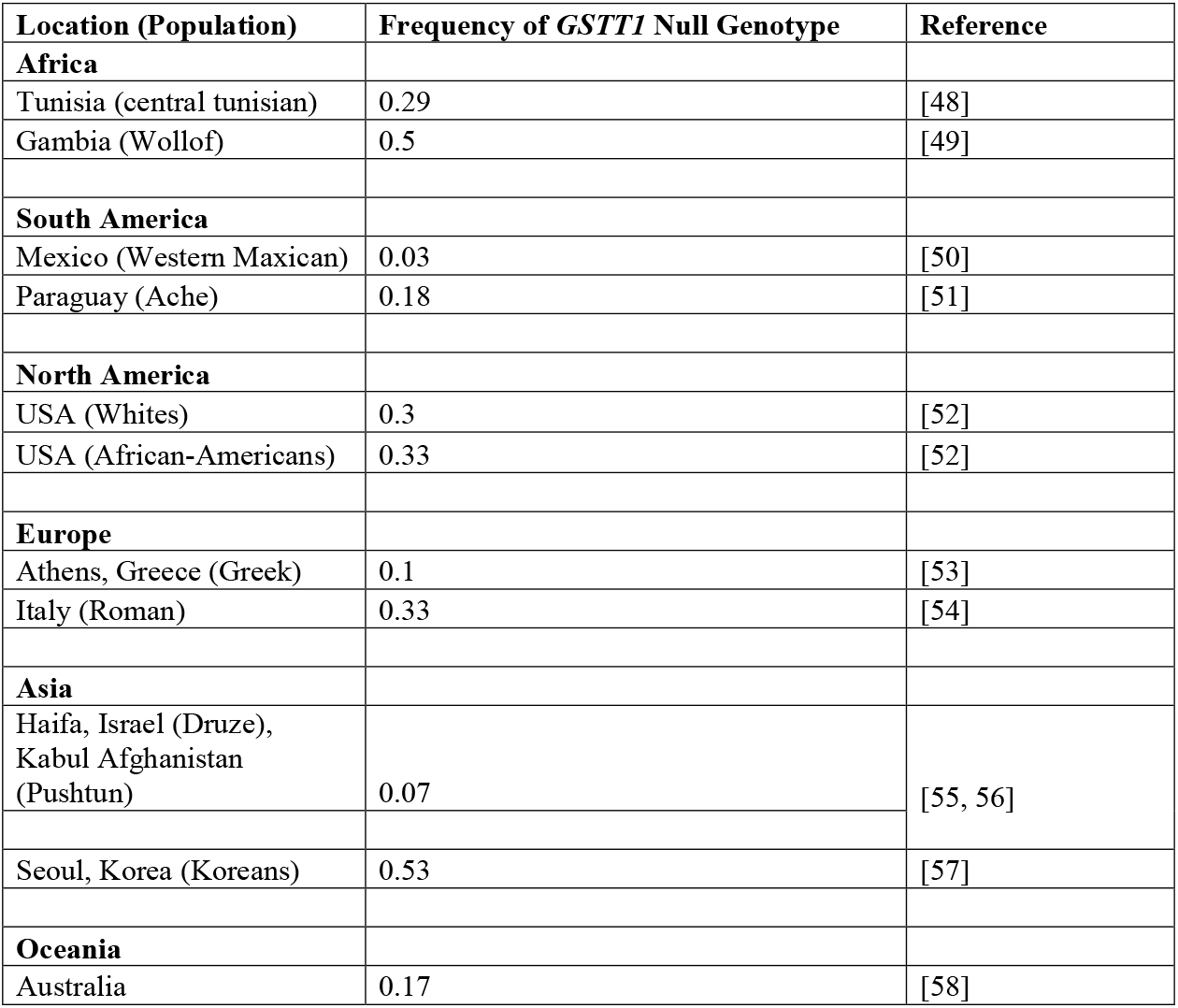
Distribution of *GSTT1*-absent genotype in different regions across the globe.

In Pakistan, the frequency of *GSTT1* null genotype in healthy individuals has been reported in the range of 0.06 – 0.24 [16-18, 27-37]. This wide range may be attributed to limited sample sizes, population admixture and differences in methodologies. Similarly, variations in the frequency of this genotype in breast cancer patients from Pakistan have been reported: 8% [16] and 27% (erroneously reported as 49% in the text) [17]. Here we report a frequency of 21%. In contrast to the studies conducted in Punjab/Central Pakistan [16, 17], the present study was carried out in Southern-Pakistan, where majority of the patients belonged to Sindhi and a self-defined Urdu-Speaking ethnicities (25% each).

Our case-control analysis is in agreement with a number of studies reported from different parts of the world [11, 59].

The present study is the only association analysis report of the *GSTT1*-absent genotype with selected clinical parameters in breast cancers from this region. It is the first report of significant association of *GSTT1*-absent genotype with decreased OS in primary breast cancer patients. An earlier study from China reported such an association with untreated metastatic breast cancers [60].

The strength of our study is the underlying unique population, where molecular data for breast cancer risk and clinical parameters is scarce. The limitations of the study are sample size and the dearth of information for known risk-factors and clinical parameters, primarily due to restriction of resources in the healthcare. The present study highlights the importance of conducting rigorous molecular epidemiology studies in order to devise evidence-based better strategies in breast cancer management, particularly in resource-limited settings.

### Conclusions

In conclusion, the significant contribution of *GSTT1*-absent genotype to the breast cancer risk in Pakistani population is reported for the first time in literature. A unique finding of this study was the association of this genotype with significantly shorter OS in breast cancer patient post standard treatment, which has not been reported earlier. These observations are biologically plausible. If validated further through multiple center studies and larger sample sizes, absence of *GSTT1* gene could serve as a risk and survival marker in breast cancers, at least in specific population.

## Data Availability

Data available on request

## Acknowledgments

We would like to thank the participants in the study..

## Compliance with ethical standards

### Funding

No funding was received for this study

### Ethical approval

The protocol was approved by the ethics review committees of the participating institutions: International Center for Chemical and Biological Sciences (ICCBS), University of Karachi, Karachi, Pakistan [ICCBS/IEC-016-BS/HT-2016/Protocol/1.0]; Atomic Energy Medical Centre (AEMC), Jinnah Postgraduate Medical Centre (JPMC), Karachi, Pakistan [Admin-3 (257)/2016]; Sindh Institute of Urology and Transplantation (SIUT), Karachi, Pakistan [18]. The study was conducted according to the Declaration of Helsinki and its further amendments.

### Participants’ consent

The participants discussed the informed consent with researchers and the voluntary enrollment in the study was documented by either signing of the written consent forms or using the thumb impressions.

## Contributions

Conceptualization: SA

Participant enrollment/Data collection: SA, SZZ, SMA, AS, MAM, AA, SF, SK

Benchwork: SA, SZZ, SMA, AS, AA, SF

Analysis: SA

Original draft: SA, SZZ, SMA

Reviewing and editing: SA

Approval of final version: all authors

